# Sexual identity and seeking mental healthcare in young adults – A national cohort study

**DOI:** 10.64898/2026.10.29.26366833

**Authors:** A. R. Khanolkar, J. Camp, A.O. Hill, V. Curcin, L. Becares

## Abstract

**Aim:** Most evidence on sexual minority health focuses on quantifying mental health inequalities with limited evidence on differences in accessing and receiving mental healthcare. This study examined associations between sexual identity and self-reported seeking and receiving mental healthcare in young adults from a population-based cohort study.

**Methods:** 12,782 individuals (22% LGBQ+) aged 23 from the UK-wide Millennium Cohort Study. Associations between sexual identity (heterosexual [reference category], mainly heterosexual, gay/lesbian and other sexual identities[LGBQ+]) assessed at age 17 and seeking mental healthcare were examined using multivariable logistic regression with sequential adjustment for sociodemographic factors (sex/ethnicity/SES), past (ages 14-17) and current (age 23) mental health, victimisation, loneliness and social support. Associations between sexual identity and receiving mental healthcare at age 23 was examined using multivariable logistic regression among those who sought help. Predicted probabilities were estimated from logistic models for interpretation.

**Results:** All LGBQ+ groups had higher odds of seeking mental healthcare than heterosexuals. In the minimally-adjusted model, predicted probability of seeking care was lowest for heterosexual (24%), highest for bisexual (45%) and gay/lesbian individuals (49%). These significant differences attenuated but persisted in fully adjusted models—narrowing the probability gap between heterosexual and LGBQ+ groups from 25% to 10%. Furthermore, mental health history independently predicted help-seeking behaviour at age 23 among heterosexuals; but not in LGBQ+ groups.

Among individuals seeking support, LGBQ+ groups had higher odds of receiving care than heterosexual peers, with highest baseline probabilities observed in the other sexual identity group (35% vs 15%). These differences remained largely unchanged after adjustment.

**Conclusion:** LGBQ+ groups face substantial disparities, demonstrating significantly higher odds of both seeking and receiving mental healthcare than heterosexual peers. Crucially, the elevated rates of receiving care persist after full adjustment, indicating that these inequalities remain largely independent of downstream mediators and mental health history.

## Introduction

Mental health conditions like depression and anxiety are the most common mental health problems and contribute significantly to global disease burden. 1.17 billion individuals were living with mental health conditions in 2023, of whom over 60% were living with major depressive or anxiety disorders(Collaborators 2026). Both major depressive and anxiety disorders are not only the most common mental health conditions across the life course but also substantially peak during adolescence and young adulthood(Collaborators 2026). This is reflected in the UK, where 1-in-5 children and young people 8-to-25 years have a mental health condition, with prevalence twice as high in women compared to men (among those aged 17-to-25 years)(Analytical Services 2023). Increasing poor mental health burden especially among young people has renewed focus on timely and appropriate access to mental healthcare services. However, in the UK only 1-in-3 individuals with mental health conditions could access support. An estimated 8 million people with mental health conditions were not in contact with mental health services, with a further 1.2 million individuals on waiting lists to access community-based mental health services(2023). There are also substantial inequalities in accessing mental healthcare services, with ethnically minoritised, lesbian, gay, bisexual and other minoritised sexual identities (LGBQ+ or sexual minority), socioeconomically disadvantaged and those with complex needs or multiple medical diagnoses facing particular barriers(2023).

LGBQ+ individuals report some of the highest levels of poor mental health and self-harming behaviours across the lifecourse, with 3-to-5 times higher rates of depression, anxiety and suicidality compared to cis-heterosexual peers(Khanolkar 2026; Khanolkar et al. 2023; Marchi et al. 2026). Limited population-based evidence exists on the patterns of access and barriers experienced by LGBQ+ groups when seeking mental healthcare, largely due to a paucity of data and a traditional focus on quantifying mental health inequities(Silveri et al. 2022). Studies have showed mixed findings with some indicating that LGBQ+ individuals are more likely to seek care compared to heterosexual individuals whereas qualitative studies have identified unique experiences encountered by LGBQ+ individuals including greater barriers in accessing services, lack of LGBQ+ relevant services, not accommodating the specific needs of the LGB+ community, longer waiting times, more negative interactions with healthcare providers, greater unmet needs and poorer treatment outcomes compared to heterosexual peers(McDermott et al. 2024; Rees et al. 2021; Rimes et al. 2019; Silveri et al. 2022). Larger quantitative UK-based surveys like the General Practice Patient Survey (GPPS) and the National LGBT Survey have showed that while overall access and primary care satisfaction remains high, LGBQ+ individuals experience longer waiting times, and those with long-term mental health conditions reported worse experiences for care involvement and provider interpersonal skills(2018; Gomes-Mendes et al. 2026). Interestingly, these individuals frequently utilized informal help-seekingresources before attempting to schedule formal appointments. However, these studies did not disaggregate sexual identity differences in seeking mental healthcare, differences by LGBQ+ subgroup, account for past mental health history and experiences of discrimination or lacked appropriate comparison groups – critical gaps in understanding patterns of access. (2018).

Despite UK government and the NHS commitments to reduce wait times and improve access among groups that face disproportionate inequalities in accessing mental healthcare, audits show that access targets for marginalized groups—including LGBQ+ groups—have not improved (2023). LGBQ+ individuals face unique barriers to mental healthcare services due to minoritised identity-related stigma and discrimination and the lack of specialised healthcare services among mainstream providers. A more comprehensive understanding of healthcare-seeking patterns is important to reduce inequalities and improve access especially among young adults who experience the highest rates of common mental health conditions like depression and anxiety. To address these gaps, this study examined (1) sexual identity differences in seeking mental healthcare including differences between LGBQ+ subgroups and (2) whether this behaviour was differentially driven by mental health history or concurrent mental health problems in early adulthood.

## Methods

### Study design and participants

The Millennium Cohort Study (MCS) is a birth cohort study following 19,519 children born in 2000-2002 with participants followed over eight sweeps (9 months, 3/5/7/11/14/17 and 23 years)(Connelly and Platt 2014). Data for this study was primarily from the ages 14(MCS 6), 17(MCS 7) and 23(MCS 8) sweeps which coincides with sexual identity development, exploration and emergence of mental health problems. Detailed information including survey design can be found at: https://cls.ucl.ac.uk/cls-studies/millennium-cohort-study/. Briefly, a nationally representative birth cohort from across the UK was recruited to the MCS and included children living in non-household situations and those not born in the UK but lived in the country at recruitment. The sampling strategy was to recruit 100% of eligible children living in geographically defined areas of residence (electoral wards prior to the 2001 Census) during the eligible period. A stratified cluster sampling design was used to ensure adequate representation of families living in disadvantaged areas and from ethnic minority groups (overall response rate was 72%). Children living in disadvantaged areas, those of ethnic minority backgrounds and growing up in the smaller nations of the UK were intentionally over-sampled. Attrition was predicted by single-parent families, lower-income occupation and educational level, Black ethnicity and male sex. The STrengthening the Reporting of OBservational studies in Epidemiology (STROBE) were followed in the reporting of this manuscript.

### Sexual identity (SI)

SI–the main exposure–was self-reported at age 17. Participants chose from one of eight options (Supplemental Table 1) and were categorised into: (1)Completely heterosexual, (2)Mainly heterosexual, (3)Bisexual, (4)Gay or lesbian and (5)Other SM identities. Due to small numbers, the other SM identities category included those who chose the options ‘other’ and ‘prefer not to say’.

### Seeking mental healthcare at age 23

Participants were asked questions on whether they sought help for mental health problems (*In the last 12 months have you sought help for a mental, nervous or emotional problem?*). Those who answered yes to seeking help, were asked a follow-up question on whether they received help (*In the last 12 months have you received help for a mental, nervous or emotional problem?*) for mental problems in the past year. Both variables were binary with the options no vs yes.

### Mental health

Mental health and self-harming behaviours were captured with self-reported and repeated measures of psychological distress, self-harm and attempted suicide assessed at ages 14, 17 and 23. At age 14, psychological distress (symptoms of depression/anxiety) was assessed using the self-reported 13-item Short Mood and Feelings Questionnaire (SMFQ) and by the 6-item Kessler psychological distress scale (K6) at ages 17 and 23. Continuous summary scores for both scales were categorised to indicate those with no psychological distress vs those having symptoms of psychological distress using validated cut-offs(Angold et al. 1995; Mewton et al. 2016). At all three ages, participants self-reported self-harm in the previous year and lifetime attempted suicide which were coded as separate binary variables (no vs yes).

We created a mental health history variable to indicate mental health problems during adolescence with the categories: No mental health problems at either age, at 1 timepoint (either age 14 or 17) and at both timepoints (both ages 14 and 17; chronic).

Mental health indicators (i.e., the three binary indicators) at age 23 were included in models separately as they were measured at the same time as seeking/receiving mental healthcare.

### Experiences of victimisation across adolescence

Victimisation was assessed at ages 14, 17 and 23 with the question: *“In the past 12 months has anyone done any of these things to you?”* and participants could choose one or more options from 1.Insulted/threatened/shouted at, 2.Experiences of physical violence, 3.Hit or used a weapon, 4.Had something stolen and 5.Been sexually assaulted. At each age we created a binary variable to indicate no experiences of victimisation vs ≥1 experiences of victimisation. We then created a cumulative victimisation variable which indicated 1.Never victimised, 2.Victimised once (either age 14 or 17) and 3.Victimised twice (at both ages 14 and 17). Victimisation at age 23 was included as a separate variable in models as it was measured concurrently with seeking/receiving mental healthcare.

### Loneliness and social support

We used the short version of the UCLA Loneliness Scale to assess loneliness(Hughes et al. 2004). Participants were categorised into “not lonely”(scores:3-5) and “lonely”(scores:6-9).

To measure social support, we used questions that asked participants whether they had individuals in their current relationships (including friends and family) who made them feel safe, secure and happy. We created a binary variable which indicated those who answered 1.Very true and partly true vs 2.Not true at all.

### Other covariates

Covariates included ethnicity, assigned sex at birth (sex), and parental income. Ethnicity was self-reported by participants at age 14. Any missing information on ethnicity was first replaced with self-reported data from age 11 and subsequently by parent reported ethnicity from the age 3 sweep. Sex was assessed at the birth sweep. For analyses, ethnicity was coded as a binary variable (White vs ethnically minoritised). Parental income was used as an indicator of socioeconomic position ascertained at age 11. Household income (Organisation for Economic Co-operation and Development UK) was categorised into equalised quintiles (where quintiles 1 and 5 represent the lowest and highest income quintiles respectively). Missing data on parental income was first replaced by data from the age 14 followed by the age 3 sweeps. Participant’s higher education attainment was a binary variable assessed at age 23; None vs having a university degree.

Complete details including the original questions, all component items of scales, questions across all sweeps, how these were operationalized for analyses (including references) and how missing data were addressed are listed in Supplemental Table 2 and Supplemental Methods.

### Multiple imputation to address missing data

There was none or low (<10%) missing data for key sociodemographic variables like sex, ethnicity and parental income. Due to attrition over follow-up and higher levels of item non-response, the proportion of missing data was higher at age 23 than earlier ages. Missing data varied between 10-30% for most variables including mental health indicators, victimisation, and health seeking behaviours, loneliness, social support and auxiliary variables. Missing data in the three sweeps (ages 14 to 23) was addressed using multiple imputation with chained equations assuming data missing at random ([MAR] and including 40 imputations). The main purpose of imputation was to address missing data in mental health indicators, bullying and victimisation and health seeking behaviours at age 23. If a study participant had data on at least one mental health indicator (psychological distress, self-harm, attempted suicide recorded at any age between 14 and 23) and victimisation (recorded at any age between 14 and 23), then the participant was included in the sample, and any missing data was imputed. The multiple imputation used for this study is explained in full in Supplemental Methods.

### Statistical analyses

All statistical analyses were conducted on the imputed dataset using Stata Statistical Software: V19. College Station, TX: StataCorp LLC.

Descriptive analysis included estimating prevalence and distribution of seeking and receiving mental health care, and mental health history by sexual identity and covariates.

We first examined associations between sexual identity and whether participants sought mental healthcare using multivariable logistic regression models with sequential adjustment as follows: Model 1: adjusted for sex, ethnicity, parental income and participant’s higher education attainment, Model 2: additionally adjusted for mental health history and victimisation history, Model 3: additionally adjusted for mental health problems at age 23 (psychological distress, self-harm and attempted suicide) and victimisation at age 23, and Model 4: additionally adjusted for loneliness and social support at age 23.

Our modelling strategy was informed by the directed acyclic graph (DAG – Supplemental Figure 1), where Model 1 estimates the total effect of sexual identity on seeking mental healthcare adjusted for a minimally sufficient adjustment set. To tentatively explore the controlled direct effect (CDE)—under the descriptive assumption of fixing intermediary and concurrent covariates to a constant level—Models 2-4 adjust for all remaining downstream nodes in the DAG (i.e., fully adjusted models to see if any residual association remains).

Next, we examined associations between sexual identity and receiving mental healthcare among those individuals who sought mental healthcare using the same sequential adjustment for covariates as described above. This set of models was restricted to those participants or ‘subpopulation’ (N=3,289) who answered yes to seeking mental healthcare using the ‘subpopulation’ command ensuring that survey design and strata information are fully preserved across imputed data.

We estimated adjusted predicted probabilities to interpret odds ratios for both outcomes across the five sexual identity categories and for models 1 and 4 (to show the change in probabilities after adjusting for covariates), using the *‘margins’* command in Stata. Both odds ratios and predicted probabilities from models 1 and 4 were visualised.

Next, we examined whether mental health history (ages 14 and 17) or concurrent mental health at age 23 differentially determined seeking mental healthcare across SI groups using multiplicative interaction terms. First, a logistic regression model was run including interaction terms between sexual identity and mental health history adjusting for all model covariates. As these interaction terms were non-significant—indicating that mental health history and seeking care associations do not potentially differ by sexual identity in this sample—no further post-estimation was performed.

Second, a logistic regression model with interactions between sexual identity and number of concurrent mental health conditions at age 23 was run. As interaction terms were significant for some groups, we explored further post-estimation. Parameter decomposition via a macro loop sequentially rotated the reference category for sexual identity, isolating the specific OR and 95% CI for the effect of concurrent mental health *within each* sexual identity group. These ORs are the likelihood of seeking help for someone with 1, 2 or 3 concurrent mental health problems compared to someone from the same sexual identity group with no mental health problems at age 23. Lastly, adjusted predicted probabilities were estimated to understand interaction terms across the 20 unique profiles (five sexual identity groups and four categories of the concurrent mental health variables) and further visualised.

#### Sensitivity analysis

We ran regression models for the associations between sexual identity and seeking care and receiving care (fully adjusted model only) in the CC sample. This was done to examine whether associations were similar between the imputed and CC samples.

## Results

### Sexual identity differences in seeking mental health care

All LGBQ+ groups had substantially higher odds of seeking mental healthcare than heterosexual individuals (Model 1: total effect; e.g., mainly heterosexual: OR 1.84 [95% CI 1.55-2.18], bisexual: OR 2.63 [2.15-3.23], gay/lesbian: OR 3.10, [2.24-4.30], Figure 1 and Supplemental Table 2). ORs were substantially attenuated in the fully adjusted Model 4. The greatest attenuation in ORs occurred among bisexual (OR 1.35, 1.07-1.70) and gay/lesbianindividuals (OR 1.76, 1.20-2.58), and associations were no longer apparent in the other-identities group (OR 0.89, 0.56-1.41). In Model 1 (Table 3), the predicted probability of seeking care was lowest for heterosexual (24%) and highest among bisexual (45%) and gay/lesbian individuals (49%). In the fully adjusted Model 4, differences in seeking mental healthcare reduced, with the difference in predicted probabilities between heterosexual and gay/lesbian groups narrowing from 25% to 10%.

**Figure 1.**
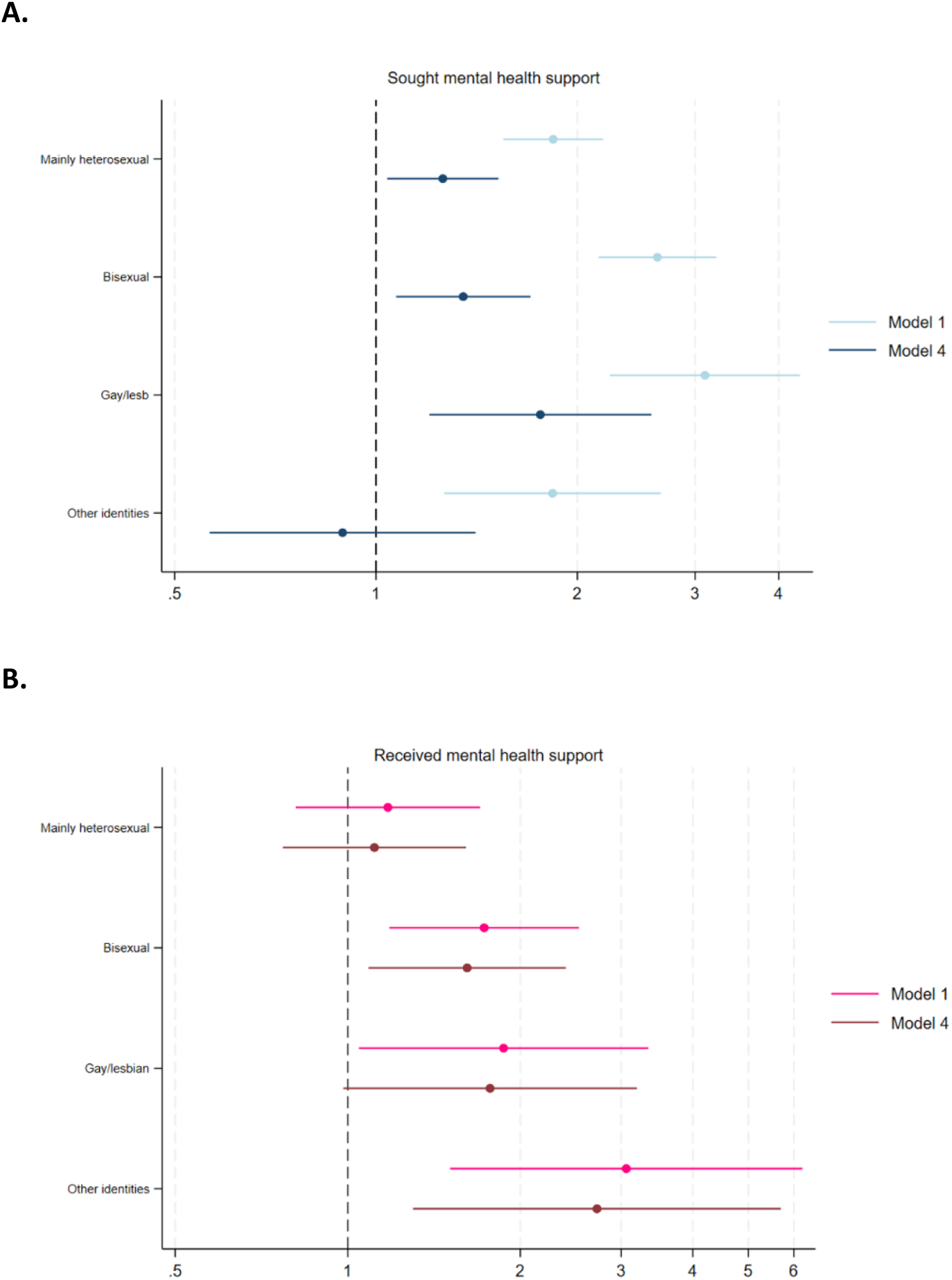
Associations between sexual identity and seeking and receiving mental healthcare in 12,782 individuals aged 23 from the Millennium Cohort Study. Note: Reference category is the heterosexual group. Estimates from logistic regression models are displayed in Supplemental Table 3. Model 1 adjusted for sex assigned at birth, ethnicity, parental income, and participant’s higher education attainment, and Model 4 additionally adjusted for mental health history and victimisation history, and mental health, victimisation, loneliness, and social support at age 23.

**Table 1.**
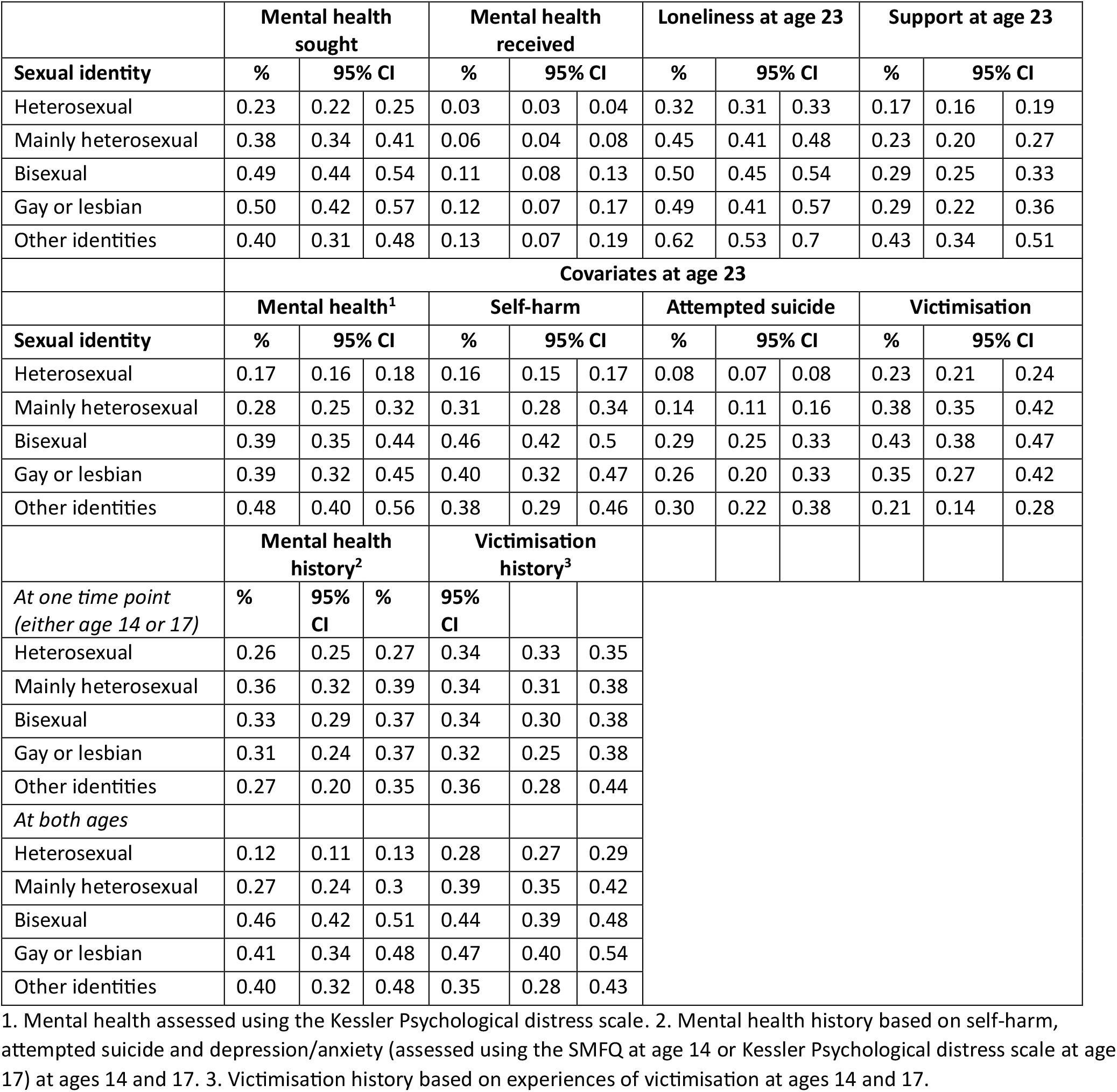
Descriptive characteristics of 12,782 individuals from the Millennium Cohort Study. Frequences (and 95% confidence intervals) are from multiply imputed data.

|  | Mental health sought |  |  | Mental health received |  |  | Loneliness at age 23 |  |  | Support at age 23 |  |  |
|---|---|---|---|---|---|---|---|---|---|---|---|---|
| **Sexual identity** | **%** | **95% CI** |  | **%** | **95% CI** |  | **%** | **95% CI** |  | **%** | **95% CI** |  |
| Heterosexual | 0.23 | 0.22 | 0.25 | 0.03 | 0.03 | 0.04 | 0.32 | 0.31 | 0.33 | 0.17 | 0.16 | 0.19 |
| Mainly heterosexual | 0.38 | 0.34 | 0.41 | 0.06 | 0.04 | 0.08 | 0.45 | 0.41 | 0.48 | 0.23 | 0.20 | 0.27 |
| Bisexual | 0.49 | 0.44 | 0.54 | 0.11 | 0.08 | 0.13 | 0.50 | 0.45 | 0.54 | 0.29 | 0.25 | 0.33 |
| Gay or lesbian | 0.50 | 0.42 | 0.57 | 0.12 | 0.07 | 0.17 | 0.49 | 0.41 | 0.57 | 0.29 | 0.22 | 0.36 |
| Other identities | 0.40 | 0.31 | 0.48 | 0.13 | 0.07 | 0.19 | 0.62 | 0.53 | 0.7 | 0.43 | 0.34 | 0.51 |
|  | **Covariates at age 23** |  |  |  |  |  |  |  |  |  |  |  |
|  | **Mental health<sup>1</sup>** |  |  | **Self-harm** |  |  | **Attempted suicide** |  |  | **Victimisation** |  |  |
| **Sexual identity** | **%** | **95% CI** |  | **%** | **95% CI** |  | **%** | **95% CI** |  | **%** | **95% CI** |  |
| Heterosexual | 0.17 | 0.16 | 0.18 | 0.16 | 0.15 | 0.17 | 0.08 | 0.07 | 0.08 | 0.23 | 0.21 | 0.24 |
| Mainly heterosexual | 0.28 | 0.25 | 0.32 | 0.31 | 0.28 | 0.34 | 0.14 | 0.11 | 0.16 | 0.38 | 0.35 | 0.42 |
| Bisexual | 0.39 | 0.35 | 0.44 | 0.46 | 0.42 | 0.5 | 0.29 | 0.25 | 0.33 | 0.43 | 0.38 | 0.47 |
| Gay or lesbian | 0.39 | 0.32 | 0.45 | 0.40 | 0.32 | 0.47 | 0.26 | 0.20 | 0.33 | 0.35 | 0.27 | 0.42 |
| Other identities | 0.48 | 0.40 | 0.56 | 0.38 | 0.29 | 0.46 | 0.30 | 0.22 | 0.38 | 0.21 | 0.14 | 0.28 |
|  | **Mental health history<sup>2</sup>** |  |  | **Victimisation history<sup>3</sup>** |  |  |  |  |  |  |  |  |
| *At one time point (either age 14 or 17)* | **%** | **95% CI** | **%** | **95% CI** |  |  |  |  |  |  |  |  |
| Heterosexual | 0.26 | 0.25 | 0.27 | 0.34 | 0.33 | 0.35 |  |  |  |  |  |  |
| Mainly heterosexual | 0.36 | 0.32 | 0.39 | 0.34 | 0.31 | 0.38 |  |  |  |  |  |  |
| Bisexual | 0.33 | 0.29 | 0.37 | 0.34 | 0.30 | 0.38 |  |  |  |  |  |  |
| Gay or lesbian | 0.31 | 0.24 | 0.37 | 0.32 | 0.25 | 0.38 |  |  |  |  |  |  |
| Other identities | 0.27 | 0.20 | 0.35 | 0.36 | 0.28 | 0.44 |  |  |  |  |  |  |
| *At both ages* |  |  |  |  |  |  |  |  |  |  |  |  |
| Heterosexual | 0.12 | 0.11 | 0.13 | 0.28 | 0.27 | 0.29 |  |  |  |  |  |  |
| Mainly heterosexual | 0.27 | 0.24 | 0.3 | 0.39 | 0.35 | 0.42 |  |  |  |  |  |  |
| Bisexual | 0.46 | 0.42 | 0.51 | 0.44 | 0.39 | 0.48 |  |  |  |  |  |  |
| Gay or lesbian | 0.41 | 0.34 | 0.48 | 0.47 | 0.40 | 0.54 |  |  |  |  |  |  |
| Other identities | 0.40 | 0.32 | 0.48 | 0.35 | 0.28 | 0.43 |  |  |  |  |  |  |

1. Mental health assessed using the Kessler Psychological distress scale. 2. Mental health history based on self-harm, attempted suicide and depression/anxiety (assessed using the SMFQ at age 14 or Kessler Psychological distress scale at age 17) at ages 14 and 17. 3. Victimisation history based on experiences of victimisation at ages 14 and 17.

**Table 2.** Predicted probabilities for seeking and receiving mental healthcare in 12,782 individuals aged 23 from the Millennium Cohort Study (visualised in Figure 2)

|  | **Seeking mental healthcare** |  |  |  |  |  |
|---|---|---|---|---|---|---|
|  | **Model 1** |  |  | **Model 4** |  |  |
| **Sexual identity** | **%** | **SE** | **95% CI** | **%** | **SE** | **95% CI** |
| Heterosexual | 0.24 | 0.01 | 0.23-0.25 | 0.27 | 0.01 | 0.25-0.28 |
| Mainly heterosexual | 0.36 | 0.02 | 0.33-0.40 | 0.30 | 0.02 | 0.27-0.34 |
| Bisexual | 0.45 | 0.02 | 0.40-0.49 | 0.32 | 0.02 | 0.28-0.36 |
| Gay/lesb | 0.49 | 0.04 | 0.41-0.56 | 0.37 | 0.04 | 0.29-0.44 |
| Other sexual identities | 0.36 | 0.04 | 0.28-0.45 | 0.25 | 0.04 | 0.18-0.32 |
|  | **Receiving mental healthcare** |  |  |  |  |  |
| Heterosexual | 0.15 | 0.01 | 0.13-0.17 | 0.14 | 0.01 | 0.12-0.16 |
| Mainly heterosexual | 0.17 | 0.02 | 0.13-0.22 | 0.16 | 0.02 | 0.11-0.20 |
| Bisexual | 0.23 | 0.03 | 0.17-0.29 | 0.21 | 0.03 | 0.15-0.27 |
| Gay/lesb | 0.25 | 0.05 | 0.15-0.35 | 0.23 | 0.05 | 0.13-0.32 |
| Other sexual identities | 0.35 | 0.08 | 0.20-0.50 | 0.31 | 0.07 | 0.16-0.46 |

Note: Models for receiving mental health care was restricted to those individuals who answered yes to seeking mental health care. Predicted probabilities are derived from models presented in Supplemental Tables 2 & 3. Model 1 adjusted for sex assigned at birth, ethnicity, parental income, and participant’s higher education attainment, and Model 4 additionally adjusted for mental health history and victimisation history, and mental health, victimisation, loneliness, and social support at age 23

**Table 3.**
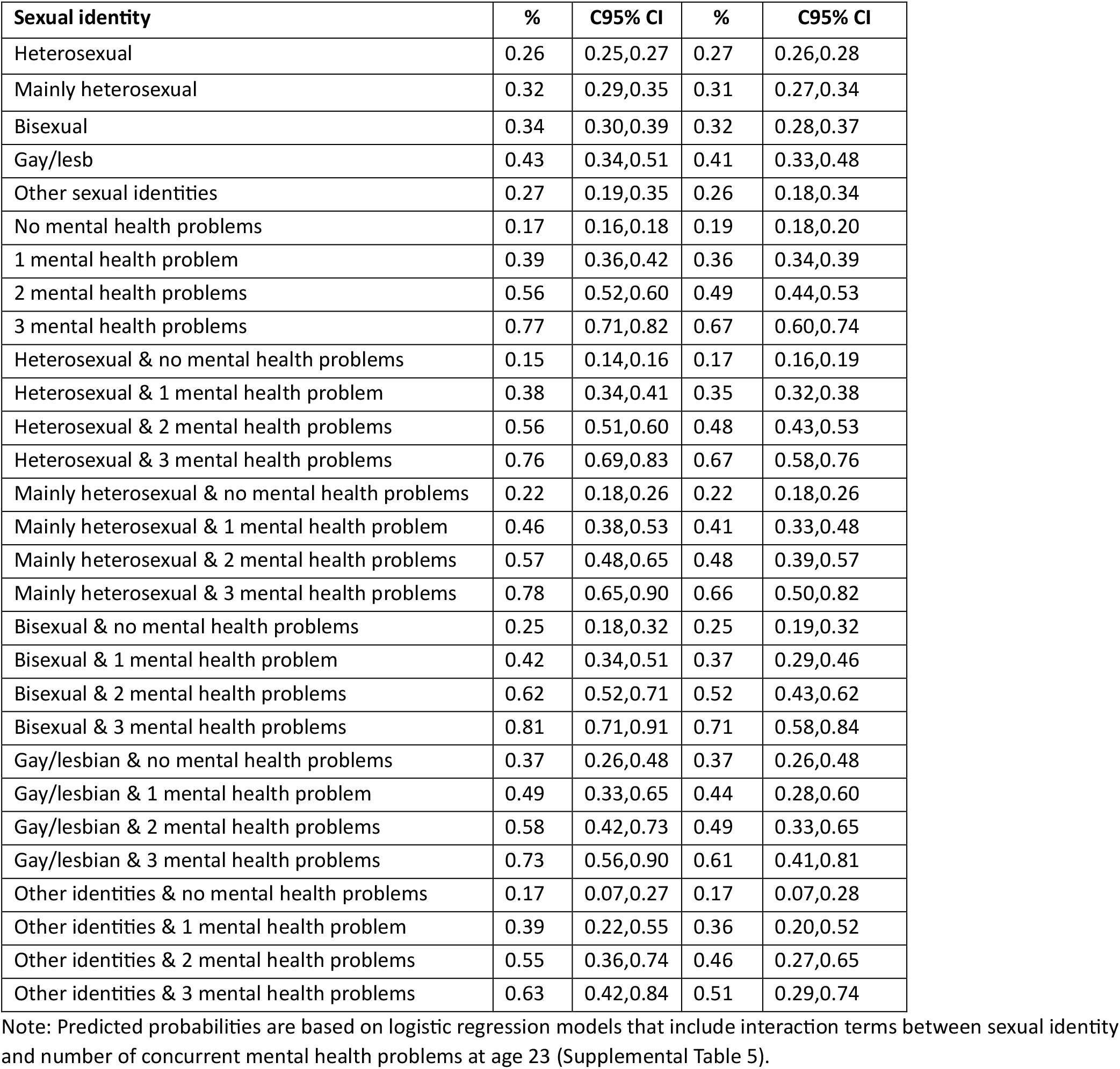
Predicted probabilities for seeking mental healthcare at age 23 based on sexual identity and number of concurrent mental health problems at age 23 (visualised in Figure 3)

| **Sexual identity** | **%** | **C95% CI** | **%** | **C95% CI** |
|---|---|---|---|---|
| Heterosexual | 0.26 | 0.25,0.27 | 0.27 | 0.26,0.28 |
| Mainly heterosexual | 0.32 | 0.29,0.35 | 0.31 | 0.27,0.34 |
| Bisexual | 0.34 | 0.30,0.39 | 0.32 | 0.28,0.37 |
| Gay/lesb | 0.43 | 0.34,0.51 | 0.41 | 0.33,0.48 |
| Other sexual identities | 0.27 | 0.19,0.35 | 0.26 | 0.18,0.34 |
| No mental health problems | 0.17 | 0.16,0.18 | 0.19 | 0.18,0.20 |
| 1 mental health problem | 0.39 | 0.36,0.42 | 0.36 | 0.34,0.39 |
| 2 mental health problems | 0.56 | 0.52,0.60 | 0.49 | 0.44,0.53 |
| 3 mental health problems | 0.77 | 0.71,0.82 | 0.67 | 0.60,0.74 |
| Heterosexual & no mental health problems | 0.15 | 0.14,0.16 | 0.17 | 0.16,0.19 |
| Heterosexual & 1 mental health problem | 0.38 | 0.34,0.41 | 0.35 | 0.32,0.38 |
| Heterosexual & 2 mental health problems | 0.56 | 0.51,0.60 | 0.48 | 0.43,0.53 |
| Heterosexual & 3 mental health problems | 0.76 | 0.69,0.83 | 0.67 | 0.58,0.76 |
| Mainly heterosexual & no mental health problems | 0.22 | 0.18,0.26 | 0.22 | 0.18,0.26 |
| Mainly heterosexual & 1 mental health problem | 0.46 | 0.38,0.53 | 0.41 | 0.33,0.48 |
| Mainly heterosexual & 2 mental health problems | 0.57 | 0.48,0.65 | 0.48 | 0.39,0.57 |
| Mainly heterosexual & 3 mental health problems | 0.78 | 0.65,0.90 | 0.66 | 0.50,0.82 |
| Bisexual & no mental health problems | 0.25 | 0.18,0.32 | 0.25 | 0.19,0.32 |
| Bisexual & 1 mental health problem | 0.42 | 0.34,0.51 | 0.37 | 0.29,0.46 |
| Bisexual & 2 mental health problems | 0.62 | 0.52,0.71 | 0.52 | 0.43,0.62 |
| Bisexual & 3 mental health problems | 0.81 | 0.71,0.91 | 0.71 | 0.58,0.84 |
| Gay/lesbian & no mental health problems | 0.37 | 0.26,0.48 | 0.37 | 0.26,0.48 |
| Gay/lesbian & 1 mental health problem | 0.49 | 0.33,0.65 | 0.44 | 0.28,0.60 |
| Gay/lesbian & 2 mental health problems | 0.58 | 0.42,0.73 | 0.49 | 0.33,0.65 |
| Gay/lesbian & 3 mental health problems | 0.73 | 0.56,0.90 | 0.61 | 0.41,0.81 |
| Other identities & no mental health problems | 0.17 | 0.07,0.27 | 0.17 | 0.07,0.28 |
| Other identities & 1 mental health problem | 0.39 | 0.22,0.55 | 0.36 | 0.20,0.52 |
| Other identities & 2 mental health problems | 0.55 | 0.36,0.74 | 0.46 | 0.27,0.65 |
| Other identities & 3 mental health problems | 0.63 | 0.42,0.84 | 0.51 | 0.29,0.74 |

Note: Predicted probabilities are based on logistic regression models that include interaction terms between sexual identity and number of concurrent mental health problems at age 23 (Supplemental Table 5).

**Figure 2.**
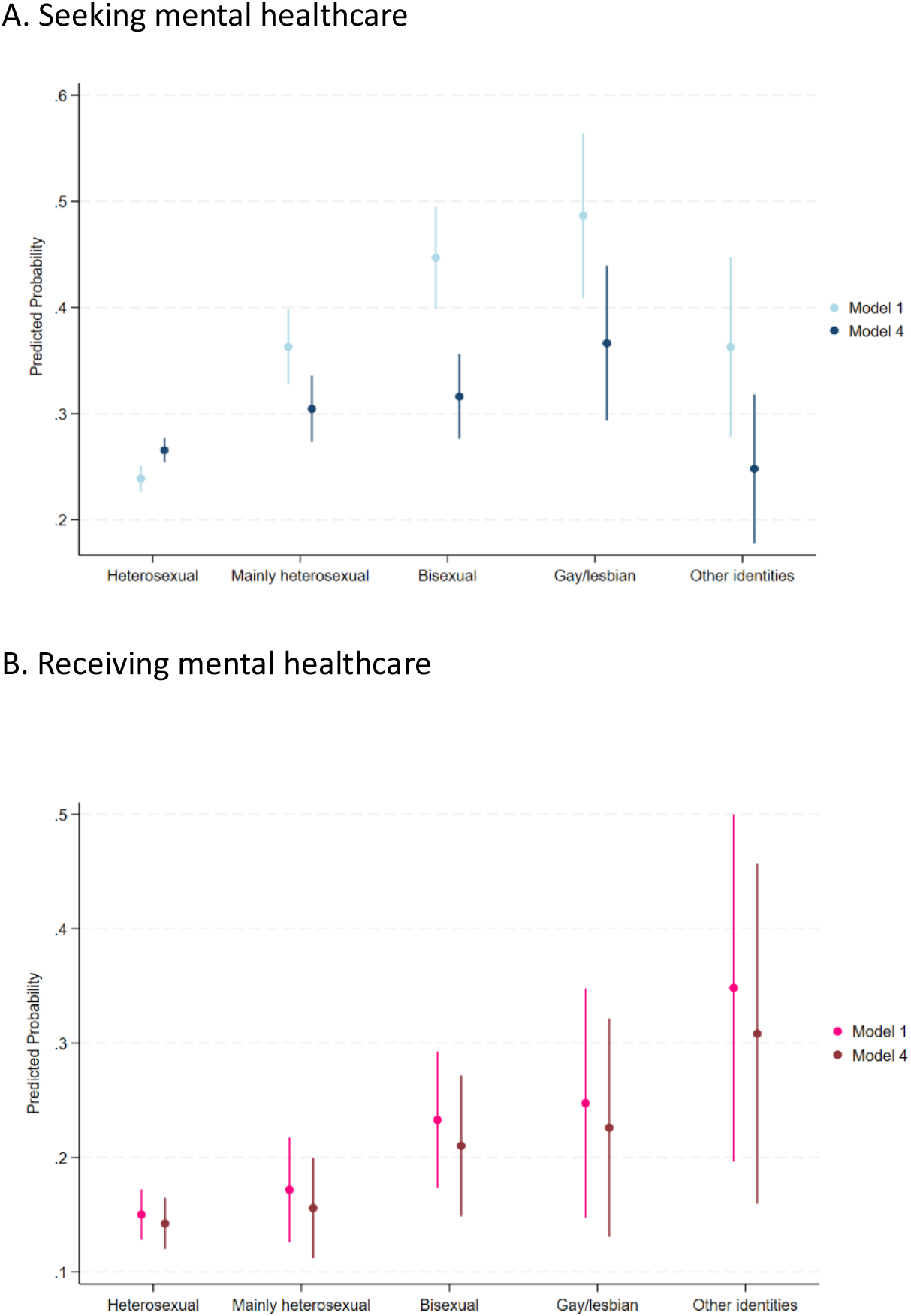
Predicted probabilities for seeking and receiving mental healthcare in 12,782 individuals aged 23 from the Millennium Cohort Study. Note: Predicted probabilities based on estimates from logistic regression models displayed in Supplemental Tables 2 & 3. Model 1 adjusted for sex assigned at birth, ethnicity, parental income, and participant’s higher education attainment, and Model 4 additionally adjusted for mental health history and victimisation history, and mental health, victimisation, loneliness, and social support at age 23.

### Mental health history and concurrent mental health as determinants of seeking mental healthcare

Among heterosexual individuals, mental health history independently increased help-seeking odds at ages 23, with a gradient, (Supplemental Table 4, Model 1: OR 1.72, 1.47-2.01 and OR 3.10, 2.58-3.73, for mental problems at either age or both ages, respectively). However, none of the sexual identity–mental health history interaction terms were statistically significant, indicating that the positive association between mental health history and later health-seeking behaviour did not reliably differ across SI groups.

**Table 4.** Associations between sexual identity, number of concurrent mental health problems and seeking mental healthcare age at 23: within sexual identity group differences.

|  | **Heterosexual** |  | **Mainly heterosexual** |  | **Bisexual** |  | **Gay/lesbian** |  | **Other sexual identities** |  |
|---|---|---|---|---|---|---|---|---|---|---|
| **Concurrent MH problems at age 23** | **OR** | **95% CI** | **OR** | **95% CI** | **OR** | **95% CI** | **OR** | **95% CI** | **OR** | **95% CI** |
| No MH problems | **Ref** |  | **Ref** |  | **Ref** |  | **Ref** |  | **Ref** |  |
| 1 MH problem | **2.74** | **2.32-3.24** | **2.54** | **1.68-3.84** | **1.83** | **1.12-2.98** | 1.36 | 0.59-3.15 | **2.76** | 1.00-7.57 |
| 2 MH problems | **4.82** | **3.82-6.08** | **3.48** | **2.22-5.44** | **3.45** | **2.02-5.92** | 1.70 | 0.76-3.83 | **4.35** | **1.52-12.43** |
| 3 MH problems | **10.86** | **7.10-16.61** | **7.73** | **3.57-16.76** | **8.19** | **3.85-17.43** | **2.86** | **1.04-7.90** | **5.45** | **1.59-18.65** |

Note:  Text in bold indicate 95% CI that do not include one. MH: mental health. Models included interaction terms between sexual identity and concurrent mental health problems, and adjusted for sex at birth, ethnicity, parental income, participant’s higher education attainment, mental health history, victimisation history, victimisation, social support and loneliness at age 23.

In the fully adjusted logistic interaction model (Supplemental Table 5 and Table 4), heterosexual individuals had a steep gradient in help-seeking odds that increased with greater number of mental health problems at age 23, e.g., OR 2.74 (2.32-3.24) to OR 4.82 (3.82-6.08) to OR 10.86 (7.10–16.61) for 1, 2 and 3 mental health problems, respectively. A significant negative interaction was observed for gay and lesbian individuals with two (OR 0.35, 0.16-0.80) and three mental health (OR 0.26, 0.09–0.81) problems, indicating a statistically less steep slope. This effect modification is displayed as adjusted predicted probabilities (Figure 3 and Table 5) where gay/lesbian individuals with no mental health problems (as measurable in this study) already have a substantially elevated baseline probability of seeking care compared to heterosexual peers (37% 0.26–0.48 vs. 17%, 0.16–0.19).

**Figure 3.**
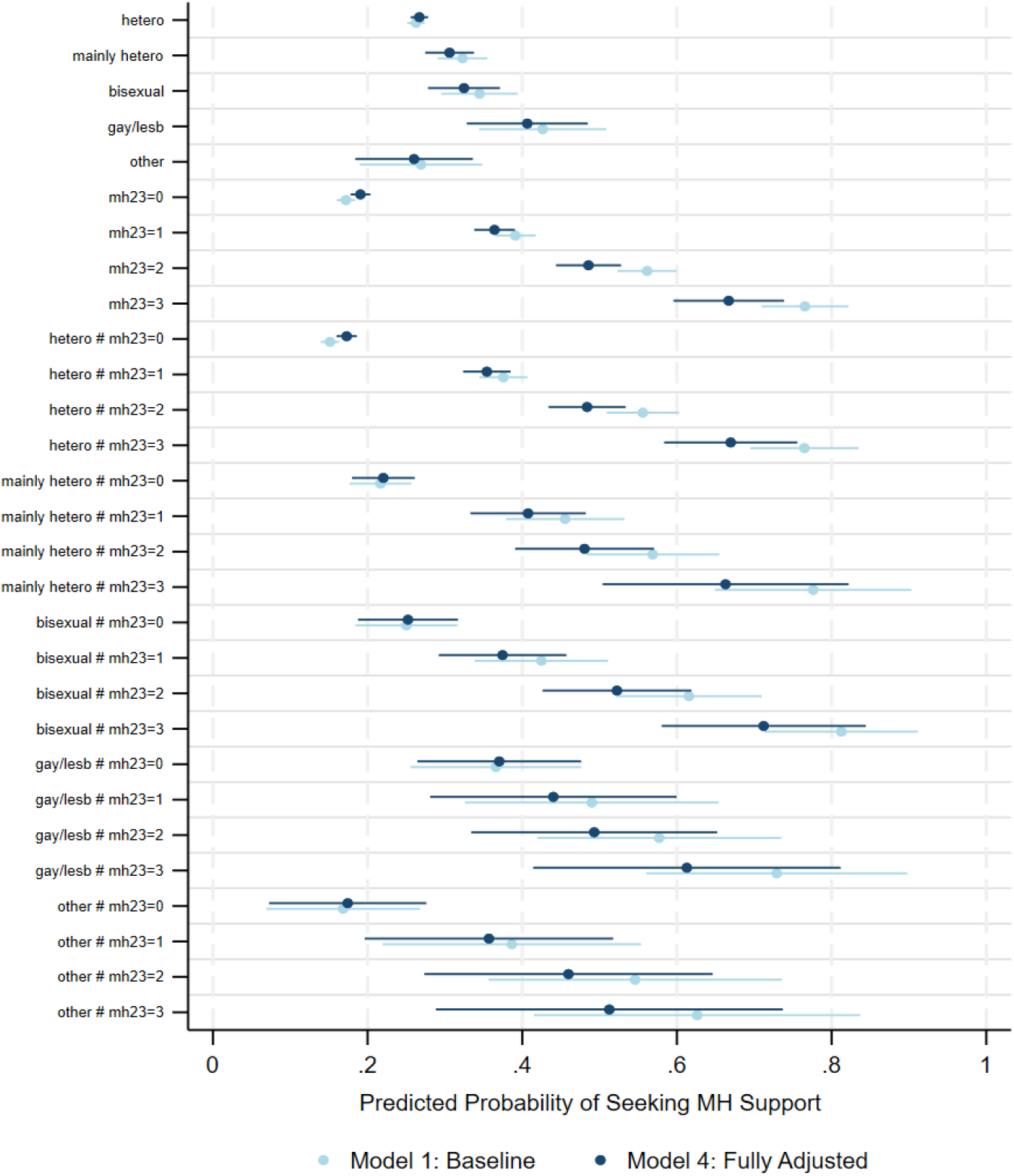
Predicted probabilities for seeking mental health care at age 23 based on sexual identity and number of concurrent mental health problems at age 23. Model 1 adjusted for sex, ethnicity, parental income, and participant’s higher education attainment. Model 2 additionally adjusted for mental health history, victimisation history, victimisation, loneliness and social support at age 23. Predicted probabilities presented in Table 3 (based on regression estimates presented in Supplemental Table Xx). Regression models included an interaction term between the sexual identity and number of concurrent mental health problems variables. MH23=mental health at age 23. MH: mental health Concurrent mental health problems include self-reported psychological distress, self-harm and attempted suicide reported at age 23.

However, for three mental health problems, these probabilities are similar for heterosexual and gay/lesbian groups (61%, 0.41–0.81 vs. 67%, 0.58–0.76). Mainly heterosexual and bisexual individuals start in the middle and climb steadily up to this same high level, while the other sexual identity group mirrors the heterosexual pattern almost exactly across all three levels of mental health problems.

Within-sexual identity group analyses revealed a gradient of increasing odds for health seeking behaviour with increasing number of concurrent mental health problems at age 23 for all SI groups. For example, having three mental health problems increased help-seeking odds approximately eight-to eleven-fold for heterosexual (OR 10.86, 7.10-16.61, Table 4), mainly heterosexual (OR 7.73, 3.57–16.76), and bisexual (OR 8.19, 3.85–17.43) groups, this gradient is substantially less pronounced for gay/lesbian individuals (OR 2.86, 1.04–7.90).

### Sexual identity differences in receiving mental health care

Among individuals who sought mental healthcare support, SM groups generally had higher odds of receiving care than heterosexual peers. In the minimally adjusted Model 1 (total effect, Supplemental Table 3), highest odds were observed for the OSI (OR 3.06 [1.51-6.21]), followed by gay/lesbian (OR 1.87 [1.05-3.34]) and bisexual (OR 1.73 [1.18-2.53]) individuals. In the fully adjusted Model 4, these odds remained significantly higher for the OSI group (OR 2.66 [1.29-5.49]) and bisexual (OR 1.62, [1.09-2.40]) groups, but less so for the gay/lesbian group (OR 1.76, [0.97-3.18]). No significant differences were observed for the mainly heterosexual group. In Model 1, the predicted probability of receiving care was lowest among heterosexual individuals (15%, Table 2) and highest for the OSI group (35%). These probabilities did not change substantially in Model 4 (ranging from 14% for heterosexual to 31% for the OSI groups), demonstrating that the substantial baseline differences in receiving care persist almost entirely independent of the downstream mediators.

## Discussion

Our study found that LGBQ+ young adults were significantly more likely to self-report seeking help for mental health conditions compared to heterosexual counterparts, and among those that did seek help, LGBQ+ groups were also more likely to receive help than heterosexuals. These findings were generally observed among all sexual minority subgroups. We found some evidence indicating that heterosexual individuals with poorer mental health in adolescence were more likely to seek care at age 23 but this association was not observed in LGBQ+ groups.

Heterosexual individuals showed a steep gradient where help-seeking odds increased substantially with the number of concurrent mental health problems at age 23, a pattern mirrored IN LGBQ+ groups. However, gay and lesbian young adults had a significantly less pronounced gradient due to a substantially elevated baseline probability of seeking care when reporting no concurrent problems.

### Strengths and limitations

Our study benefits from a large sample drawn from a nationally representative and contemporary cohort, making findings generalisable to young adults in the UK. A relatively large proportion (22%) of MCS participants identified as LGBQ+ which provided sufficiently large sample sizes to enable analysis across sexual minority subgroups.

Sexual identity and ethnicity were self-reported, which is the gold-standard. Further, the MCS captures sexual identity before our mental health outcomes at age 23, ensuring temporality. We accounted for loneliness and lack of social support which are more common among LGBQ+ young adults than heterosexual counterpart and also associated with poorer mental health but often excluded in studies on sexual minority health.

Like most birth cohorts, the MCS has experienced significant attrition over 23 years but participants attending the ages 17 and 23 assessments remain representative of the original sample recruited at birth(Fitzsimons 2020). We addressed missing data (item nonresponse) using multiple imputation further strengthened with a rich set of auxiliary variables measured at several timepoints increasing the validity and robustness of the imputation model(Sterne et al. 2009; White et al. 2011). The mental history variable measures the most common mental health problems (depression and anxiety) using the validated and commonly used SMFQ and Kessler scales(Angold et al. 1995), but we acknowledge that it does not account for all possible mental health conditions in adolescence. We also acknowledge that sexuality is fluid and may change across adolescence and early adulthood which we were unable to account for. These analyses capture associations between sexual identity and mental healthcare at a critical period in the life course, with important implications for mental health outcomes regardless of sexual orientation in later stages of the life course. Like most national surveys, the MCS does not capture sexual minority relevant information including familial, peer or community-level support, which can help reduce mental health problems and also moderate pathways to seeking mental healthcare. Despite a large sample size of 12,800 individuals, models including interaction terms resulted in estimates with relatively wide confidence intervals for the gay/lesbian and other sexual identity groups and we advise caution in interpretation of observed associations. Our measure of ‘receiving care’ is broad and does not account for important features like timeliness, adequacy, intensity and quality of care received, and was only measured among those individuals who had already sought help.

To our knowledge, this is the first study in the UK to report these findings using a nationally representative sample, longitudinal in design and report differences across sexual minority subgroups. Our study confirms previous findings from the limited but growing evidence base that sexual minority groups are more likely to seek help for mental health conditions than heterosexual peers(Cochran et al. 2017; Conner 2023; Cronin et al. 2021; Dunbar et al. 2017; Filice and Meyer 2018; Platt et al. 2018; Roxo et al. 2026). However, most studies examining mental health service use by sexual identity are based on non-probability or convenience samples often recruited online, focus on specific groups like college students, examine all sexual minority subgroups together, or are cross-sectional in design. Further, most studies originate from the USA where healthcare system access is fundamentally different from the UK and wider Europe(Filice and Meyer 2018; Roxo et al. 2026). The most recent European population-based study focused on young adults in Sweden and found that sexual minority individuals reported higher rates of mental health service use(Roxo et al. 2026). However, this study was smaller, examined all sexual minority groups together and did not account for past mental health problems, victimisation and loneliness. Some studies have reported differences in access rates by sexual minority subgroup and gender(Filice and Meyer 2018; Platt et al. 2018; Roxo et al. 2026). We found higher rates of seeking help among bisexual and gay/lesbian groups compared to mainly heterosexual individuals, but differences were relatively small.

A key contribution of our study is accounting for past and current mental health conditions in examining associations between sexual identity and seeking/receiving care. To our knowledge, only one large population-based study found that adjusting for past mental health problems attenuated associations in lesbian, gay and bisexual groups and associations were no longer evident in the other sexual identities, like our findings(Platt et al. 2018). However, our study builds on this by using more defined measures of past and concurrent mental health and testing associations using appropriate interactions. However, we acknowledge that past and concurrent mental health are more likely intermediaries in the associations between sexual identity and seeking/receiving care as described in our DAG. Our study also confirms previous findings that LGBQ+ groups are more likely to receive mental healthcare, but comparisons are limited as receiving care is defined or measured differently across studies.

Our findings provide evidence for the persistent and substantially higher burden of poor mental health experienced by LGBQ+ young adults. Previous studies, including the MCS, consistently show that LGBQ+ groups experience significantly worse mental health, victimisation and adverse childhood experiences across adolescence and early adulthood than heterosexual peers(Chandrasekar et al. 2024; Khanolkar and Becares 2025; Khanolkar et al. 2023). In this regard, the higher odds for seeking and receiving care rather reflects this systemic higher burden rather than absolute greater rates of seeking and receiving healthcare. This is also reflected in our findings of number of increasing concurrent mental health conditions at age 23 were associated with increasing probability of seeking care with a stark gradient in the full sample and among heterosexuals, but this gradient while evident in LGBQ+ groups was substantially smaller in gay/lesbian individuals. Further, our findings should be interpreted in the light of several studies that have found that despite higher rates of seeking care among LGBQ+ groups than heterosexual peers, the former continue to experience substantially higher rates of unmet care needs(Roxo et al. 2026; Williams and Chapman 2011).

Overall, our study shows that, compared to heterosexual young adults, LGBQ+ young individuals are more likely to report seeking help for mental health problems. Among those who sought help, sexual minority individuals were also more likely to receive help than heterosexual peers. Associations between mental health history in adolescence and seeking mental healthcare at age 23 did not differ by sexual identity. However, concurrent mental health problems were associated with greater odds for seeking care with a clear gradient among heterosexuals but less so among LGB+ groups. Future studies need to explore the broader remit of healthcare access including timeliness, appropriateness, unmet needs and other LGB+ specific barriers.

## Required Statements

## Acknowledgements

The Economic and Social Research Council funds the Centre for Longitudinal Studies (CLS) Resource Centre (ES/W013142/1) which provides core support for the CLS cohort studies. While the CLS Resource Centre makes these data available, CLS does not bear any responsibility for the analysis or interpretation of these data by researchers. The CLS cohorts are only possible due to the commitment and enthusiasm of their participants; their time and contribution are gratefully acknowledged.

## Data availability

The data used in this study is from the Millennium Cohort Study, and the data can be downloaded via special license and after registering on the UK Data Service website (https://ukdataservice.ac.uk/) and agreeing to all terms and conditions. All data used in this study are completely anonymized and participants cannot be identified. Further information on the Millennium Cohort Study can be found on the study’s website: https://cls.ucl.ac.uk/cls-studies/millennium-cohort-study/

## Ethical Standards

Ethics approval for the MCS was obtained from the National Research Ethics Service Committee London— Central (reference 13/LO/1786). Informed parental consent was obtained in advance of data collection up to age 14 years. Cohort members ≥16 years provided verbal consent to take part in the study. Participants were able to refuse to participate in any element of data collection or withdraw from the study at any time. This study did not require ethical approval as it is a secondary data analysis of de-identified data protecting the identity of participants. This study conformed to the principles of the Helsinki Declaration.

## References

(2018) National LGBT Survey. London. Available at https://assets.publishing.service.gov.uk/media/5b3cb6b6ed915d39fd5f14df/GEO-LGBT-Survey-Report.pdf (accessed.

(2023) Progress in improving mental health services in England. London. Available at https://www.nao.org.uk/wp-content/uploads/2023/02/Progress-in-improving-mental-health-services-CS.pdf (accessed.

Analytical Services PH, Clinical Audit & Specialist Care Team (2023) Mental Health of Children and Young People in England 2023 – follow up to the 2017 survey. Available at https://digital.nhs.uk/data-and-information/publications/statistical/mental-health-of-children-and-young-people-in-england/2023-wave-4-follow-up (accessed.

Angold A, Costello EJ, Messer SC, Pickles A, Winder F and Silver D (1995) Development of a short questionnaire for use in epidemiological studies of depression in children and adolescents. International Journal of Methods in Psychiatric Research 5(4), 237–249.

Chandrasekar R, Karamanos A, Learoyd A and Khanolkar AR (2024) Sexual identity-related inequalities in associations between adverse childhood experiences and health in late adolescence-A national cohort study. PLoS One 19(12), e0312161. 10.1371/journal.pone.0312161.

Cochran SD, Bjorkenstam C and Mays VM (2017) Sexual orientation differences in functional limitations, disability, and mental health services use: Results from the 2013-2014 National Health Interview Survey. J Consult Clin Psychol 85(12), 1111–1121. 10.1037/ccp0000243.

Collaborators GBDMD (2026) Updated trends in the global prevalence and burden of mental disorders, 1990-2023: a systematic analysis for the Global Burden of Disease Study 2023. Lancet 407(10543), 2040–2064. 10.1016/S0140-6736(26)00519-2.

Connelly R and Platt L (2014) Cohort profile: UK Millennium Cohort Study (MCS). Int J Epidemiol 43(6), 1719–1725. 10.1093/ije/dyu001.

Conner Ckl, K.M.; Dermody, S.S. (2023) Access and barriers to health services among sexual and gender minority college students. Psychology of Sexual Orientation and Gender Diversity 10(3), 498–508. 10.1037/sgd0000559.

Cronin TJ, Pepping CA, Halford WK and Lyons A (2021) Mental health help-seeking and barriers to service access among lesbian, gay, and bisexual Australians. Australian Psychologist 56(1), 46–60. 10.1080/00050067.2021.1890981.

Dunbar MS, Sontag-Padilla L, Ramchand R, Seelam R and Stein BD (2017) Mental Health Service Utilization Among Lesbian, Gay, Bisexual, and Questioning or Queer College Students. J Adolesc Health 61(3), 294–301. 10.1016/j.jadohealth.2017.03.008.

Filice E and Meyer SB (2018) Patterns, predictors, and outcomes of mental health service utilization among lesbians, gay men, and bisexuals: A scoping review. Journal of Gay & Lesbian Mental Health 22(2), 162–195. 10.1080/19359705.2017.1418468.

Fitzsimons Eh, L..; Smith, K.: Gilbert, E.: Calderwood, L.: AgaliotiSgompou, V.: Veeravalli, S., Silverwood, R.: Ploubidis, G. (2020) Millennium Cohort Study Age 17 Sweep (MCS7): User guide. London.

Gomes-Mendes A, McManus S, van der Ven E, Dima D and Fett AK (2026) Primary care experiences at the intersection of sexual minority status and long-term mental health conditions in England: a cross-sectional analysis using the English General Practice Patient Survey. EClinicalMedicine 96, 104016. 10.1016/j.eclinm.2026.104016.

Hughes ME, Waite LJ, Hawkley LC and Cacioppo JT (2004) A Short Scale for Measuring Loneliness in Large Surveys: Results From Two Population-Based Studies. Res Aging 26(6), 655–672. 10.1177/0164027504268574.

Khanolkar AR (2026) Health inequalities among non-lesbian, gay and bisexual sexual minority adolescents: Findings from a national UK cross-sectional study. BMC Glob Public Health 4(1). 10.1186/s44263-026-00286-7.

Khanolkar AR and Becares L (2025) Prevalence of past homelessness, mental health and health risk behaviours among sexual minority young people in the UK: insights from the Millennium Cohort Study. BMC Public Health 25(1), 4241. 10.1186/s12889-025-25429-7.

Khanolkar AR, Frost DM, Tabor E, Redclift V, Amos R and Patalay P (2023) Ethnic and Sexual Identity-Related Inequalities in Adolescent Health and Well-Being in a National Population-Based Study. LGBT Health 10(1), 26–40. 10.1089/lgbt.2021.0473.

Marchi M, Ferrazzi G, Pollice G, Pingani L, Ferrari S, Ventriglio A and Galeazzi GM (2026) Mental Health in Sexual and Gender Minority Populations: A Systematic Review of Systematic Reviews With Narrative Synthesis and a Network Meta-Analysis of Suicide Attempts. Int J Soc Psychiatry 72(2), 205–218. 10.1177/00207640251367069.

McDermott E, Eastham R, Hughes E, Johnson K, Davis S, Pryjmachuk S, Mateus C, McNulty F and Jenzen O (2024) “What Works” to Support LGBTQ+ Young People’s Mental Health: An Intersectional Youth Rights Approach. Int J Soc Determinants Health Health Serv 54(2), 108–120. 10.1177/27551938241230766.

Mewton L, Kessler RC, Slade T, Hobbs MJ, Brownhill L, Birrell L, Tonks Z, Teesson M, Newton N, Chapman C, Allsop S, Hides L, McBride N and Andrews G (2016) The psychometric properties of the Kessler Psychological Distress Scale (K6) in a general population sample of adolescents. Psychol Assess 28(10), 1232–1242. 10.1037/pas0000239.

Platt LF, Wolf JK and Scheitle CP (2018) Patterns of Mental Health Care Utilization Among Sexual Orientation Minority Groups. J Homosex 65(2), 135–153. 10.1080/00918369.2017.1311552.

Rees SN, Crowe M and Harris S (2021) The lesbian, gay, bisexual and transgender communities’ mental health care needs and experiences of mental health services: An integrative review of qualitative studies. J Psychiatr Ment Health Nurs 28(4), 578–589. 10.1111/jpm.12720.

Rimes KA, Ion D, Wingrove J and Carter B (2019) Sexual orientation differences in psychological treatment outcomes for depression and anxiety: National cohort study. J Consult Clin Psychol 87(7), 577–589. 10.1037/ccp0000416.

Roxo L, Pachankis J and Branstrom R (2026) Sexual orientation differences in mental health service use and unmet mental health care needs: a cross-sectional population-based study of young adults. Soc Psychiatry Psychiatr Epidemiol 61(2), 225–234. 10.1007/s00127-025-02866-8.

Silveri G, Schimmenti S, Prina E, Gios L, Mirandola M, Converti M, Fiorillo A, Pinna F, Ventriglio A, Galeazzi GM, Sherriff N, Zeeman L, Amaddeo F and Italian Working Group on LMH (2022) Barriers in care pathways and unmet mental health needs in LGBTIQ + communities. Int Rev Psychiatry 34(3-4), 215–229. 10.1080/09540261.2022.2075256.

Sterne JA, White IR, Carlin JB, Spratt M, Royston P, Kenward MG, Wood AM and Carpenter JR (2009) Multiple imputation for missing data in epidemiological and clinical research: potential and pitfalls. BMJ 338, b2393. 10.1136/bmj.b2393.

White IR, Royston P and Wood AM (2011) Multiple imputation using chained equations: Issues and guidance for practice. Stat Med 30(4), 377–399. 10.1002/sim.4067.

Williams KA and Chapman MV (2011) Comparing health and mental health needs, service use, and barriers to services among sexual minority youths and their peers. Health Soc Work 36(3), 197–206. 10.1093/hsw/36.3.197.

